# Experiences of Difficulty in Hematopoietic Cell Transplant Nurses: A Qualitative Systematic Review Protocol

**DOI:** 10.1101/2025.03.04.25323395

**Authors:** Yuko Kurahashi, Satoko Okuyama

## Abstract

**Objective:** This review aimed to explore the experiences of nurses working in the field of hematopoietic stem cell transplantation (HSCT) and identify the psychological, emotional, and ethical challenges they face. Given the limited literature specifically focusing on HSCT nurses, this review also includes the experiences of nurses caring for patients with hematologic malignancies in order to provide a comprehensive evaluation of the unique difficulties and coping strategies associated with HSCT.

**Introduction:** Nursing in HSCT requires a high level of specialization and is accompanied by multifaceted challenges, including psychological stress, emotional exhaustion, and ethical dilemmas. HSCT involves destroying the patient’s bone marrow function through intensive chemotherapy and total body irradiation, followed by transplantation of hematopoietic stem cells to restore blood cell production (Kanda, 2015). Introduced in Japan in 1974, more than 5,500 procedures are performed annually. However, life-threatening risks such as graft-versus-host disease (GVHD) and severe infections remain high (Japan Data Center for Hematopoietic Cell Transplantation, 2023).

Nurses face not only technical challenges such as infection prevention and complex treatment management, but also the psychological and emotional burden of supporting patients suffering from severe stress. Many patients struggle with anxiety about relapse, isolation in sterile environments, and long-term psychological burden of recovery. The five-year survival rate for acute myeloid leukemia remains between 44% and 56% (Japan Data Center for Hematopoietic Cell Transplantation, 2023), further exacerbating the difficulties of nursing care due to the severity and complexity of the treatment.

This review examines the challenges faced by nurses in HSCT and hematologic oncology settings using qualitative and mixed methods research to clarify the realities of these experiences. In doing so, it aims to provide insights that can serve as a foundation for future support measures and educational programs.

**Inclusion Criteria:** This review included qualitative and mixed-methods studies focusing on the psychological, emotional, and ethical challenges faced by nurses caring for HSCT and hematologic malignancy patients. Studies targeting pediatric patients and those focusing on professions other than nursing were also excluded.

**Methods:** The primary search engines for this review were PubMed, CINAHL, and Ichushi-Web, and searches were conducted between May and July 2025. The search was limited to the English and Japanese literature. The study selection will involve screening titles and abstracts, followed by a full-text review of relevant studies. Methodological rigor will be critically appraised, and data will be synthesized to identify recurring themes and patterns. The reliability of the findings was evaluated based on the consistency and quality across studies.

**PROSPERO Registration Number:** CRD42024605711

## Introduction

### Rationale for the Review

Hematopoietic stem cell transplantation (HSCT) nursing is a highly specialized field that presents numerous challenges. HSCT is a treatment in which a patient’s bone marrow is destroyed through intensive chemotherapy and total body irradiation, followed by transplantation of hematopoietic stem cells derived from the bone marrow, peripheral blood, or umbilical cord blood to restore hematopoiesis (Kanda, 2015). HSCT was first introduced in 1974 in Japan, and currently, more than 5,500 procedures are performed annually (Japan Data Center for Hematopoietic Cell Transplantation, 2023).

However, HSCT carries life-threatening complications, such as graft-versus-host disease (GVHD) and severe infections. The five-year survival rate for acute myeloid leukemia patients remains challenging (44–56%), depending on the donor type and stem cell source (Japan Data Center for Hematopoietic Cell Transplantation, 2023). Post-transplant patients face fatal risks from complications (Ishida et al., 2002) and continue to suffer psychological distress due to fear of relapse, even after the acute phase (Ishida et al., 2005).

Patients undergoing rigorous treatment require strong support from nurses with specialized knowledge (Takahashi & Onishi, 2007). For example, post-transplant infection prevention requires strict dietary guidelines (Japan Society for Hematopoietic Cell Transplantation, 2017), which nurses often find challenging to explain to their patients. Additionally, patients in isolation rooms may exhibit various psychological states such as anger or emotional numbness (Akaho, 2007), making psychological support a challenge for nurses.

Under these circumstances, transplant nurses must manage not only technical challenges such as infection control and complex treatment planning, but also psychological and ethical dilemmas. In particular, nurses experience a significant burden when providing end-of-life care and supporting patients’ decision-making autonomy. HSCT nurses must possess advanced clinical knowledge and the ability to provide emotional support.

### Justification for Inclusion Criteria

This review focuses on qualitative and mixed-methods studies that examine the psychological, emotional, and ethical challenges in HSCT and hematologic oncology nursing, with the aim of clarifying the complexity of nurses’ experiences and perceptions. Studies focusing on pediatric patients were excluded because their challenges and contexts differ significantly from those of adult patients.

The inclusion of hematologic oncology nursing is justified because of the limited number of studies specifically addressing HSCT nursing. By integrating broader findings, this review aimed to highlight the unique difficulties and coping strategies specific to HSCT nursing.

### Regional Differences in Understanding

The challenges and ethical dilemmas faced by nurses may vary owing to cultural and healthcare system differences. For example, decision-making support in end-of-life care is influenced by cultural values regarding life and death as well as medical ethics. This review aimed to integrate findings that reflect diverse backgrounds while considering regional differences.

### Key Terms and Their Operational Definitions

#### Hematopoietic Stem Cell Transplantation (HSCT)

Definition: Treatment that restores bone marrow function by transplanting hematopoietic stem cells derived from bone marrow, peripheral blood, or umbilical cord blood (Kanda, 2015).

Operational Definition: In this review, HSCT specifically refers to allogeneic hematopoietic stem cell transplantation for hematologic malignancies and related disorders.

#### Hematologic Malignancies

Definition: A general term for cancers affecting the blood, bone marrow, and lymphatic system, including leukemia, lymphoma, and multiple myeloma.

Operational Definition: In this review, “hematologic malignancies” refer to a broad category of blood cancers regardless of whether HSCT is required.

#### Difficulties

Definition: A multifaceted issue in nursing practice encompasses psychological stress, emotional exhaustion and ethical dilemmas.

Operational Definition: In this review, “difficulties” refer to the psychological, emotional, and ethical burdens and challenges experienced by nurses in HSCT or hematologic oncology settings.

#### Review of Previous Studies

A preliminary search of PROSPERO, MEDLINE, the Cochrane Database of Systematic Reviews, and JBI Evidence Synthesis found no ongoing or planned systematic reviews on this topic. This review aimed to differentiate itself from previous studies by providing a systematic synthesis of psychological, emotional, and ethical challenges in HSCT and hematologic oncology nursing.

#### Objective of the Review

The objective of this review is to identify the psychological, emotional, and ethical challenges faced by nurses in HSCT and hematologic oncology nursing and to synthesize the unique difficulties and coping strategies specific to HSCT nursing.

## Review question(s)

### The question of this review is

What are the experiences of psychological, emotional, and ethical challenges among HSCT and hematologic oncology nurses?

### Sub-questions

#### 1. Specific nature of challenges

How can the psychological stress and emotional burden experienced by nurses be categorized in terms of contributing factors and specific aspects?

#### 2. Coping strategies and support

How do nurses cope with these challenges, and what types of support systems are available to assist them?

## Inclusion criteria

### Participants

This review includes nurses involved in hematopoietic stem cell transplantation (HSCT). Additionally, since research on HSCT nursing is limited, nurses who care for patients with hematologic malignancies will be included. However, studies focusing primarily on nurses caring for pediatric patients or healthcare professionals other than nurses were excluded.

### Phenomena of interest

This review considers studies that explore the psychological, emotional, and ethical challenges experienced by nurses in HSCT and hematologic oncology settings. Specifically, it includes studies that discuss how nurses cope with these challenges and the types of support needed in clinical practice.

### Context

This review focuses on the challenges experienced by HSCT and hematologic oncology nursing. The relevant contexts include nursing practice in acute care hospitals and specialized transplant centers. Geographically, this review will consider multicultural healthcare settings, including Japan, taking into account how cultural and social backgrounds influence nurses’ experiences.

### Types of studies

This review considers studies that have focused on qualitative data. Specifically, it includes studies that explore the experiences of nurses caring for HSCT and hematologic oncology patients. The review will consider, but is not limited to, the following research design:

- Phenomenology
- Grounded theory
- Ethnography
- Action research

Additionally, this review will include interpretive studies that analyze the psychological, emotional, and ethical challenges faced by nurses, as well as critical studies that examine these challenges from a broader analytical perspective.

## Methods

This systematic review will be conducted in accordance with the methodology of systematic reviews of qualitative evidence provided by the JBI. The design and implementation of this review were guided by the JBI Manual for Evidence Synthesis.

Additionally, this review was registered with PROSPERO under registration number CRD42024605711. The review will be carried out following the registered protocol and JBI methodology to ensure transparency.

If any deviations from the protocol occur during the course of the review, the reasons and details are reported and justified in this section.

### Search strategy

This review employed a comprehensive three-step search strategy to locate both published and unpublished studies.

#### 1. Initial Search

An initial limited search was conducted using MEDLINE (PubMed), CINAHL (EBSCO), and Ichushi Web. This search aimed to identify relevant literature on the topic. The text words found in the titles and abstracts of these articles as well as the index terms used to describe them were extracted. This information forms the basis for developing a comprehensive search strategy tailored to each database and information source (see Appendix # for further details).

#### 2. Comprehensive Search

A comprehensive search was conducted across each database using the keywords and index terms identified in the initial search. The databases used were MEDLINE, CINAHL, and Ichushi Web. The search strategy was adapted as needed to fit the specific characteristics of each database or information source.

#### 3. Screening Reference Lists

The reference lists of all the included articles were screened to identify additional relevant studies. This process involves examining the titles and abstracts of the references to determine their eligibility for inclusion in the review.

References from systematic reviews and related studies on the same or similar topics are also reviewed.

### Sources of Information

The following sources of information will be searched:

- **Databases:** PubMed (MEDLINE), EBSCO (CINAHL), Ichushi Web
- **Unpublished Studies:** Conference proceedings, relevant websites, and direct contact with researchers.

### Language and Time Frame

This review will include studies published in any language, and no timeframe restrictions will be applied.

### Validation and Transparency

The search strategy uses tools and filters to identify keywords and index terms. A peer review of the search strategy was conducted when necessary to ensure accuracy and transparency. Detailed descriptions of the search methods and processes are provided in the Appendix.

### Study selection

All identified citations from the search were collated and uploaded into EndNote 21 (Clarivate), a bibliographic management software, and duplicates were removed. Following a pilot test to evaluate the applicability of the screening criteria, the titles and abstracts were screened by two or more independent reviewers. Studies deemed potentially relevant were retrieved in full, and their citation details were imported into the JBI System for Unified Management, Assessment, and Review of Information (JBI SUMARI).

The full text of the selected citations will be thoroughly assessed against the inclusion criteria by two or more independent reviewers. Reasons for the exclusion of studies at the full-text stage that do not meet the inclusion criteria will be recorded and reported in the systematic review. Any disagreements between the reviewers at any stage of the selection process will be resolved through discussion or with the involvement of additional reviewers.

The search results and study selection process will be reported in the final systematic review and presented in a flow diagram based on the Preferred Reporting Items for Systematic Reviews and Meta-Analyses (PRISMA) statement.

### Assessment of methodological quality

Eligible studies were critically appraised by at least two independent reviewers using the JBI Critical Appraisal Checklist for Qualitative Research. Appraisal assesses the following ten domains:

1. Is there congruity between the stated philosophical perspective and the research methodology?
2. Is there congruity between the research methodology and the research question or objectives?
3. Is there congruity between the research methodology and the methods used to collect data?
4. Is there congruity between the research methodology and the representation and analysis of data?
5. Is there congruity between the research methodology and the interpretation of results?
6. Is there a statement locating the researcher culturally or theoretically?
7. Is the influence of the researcher on the research, and vice-versa, addressed?
8. Are participants, and their voices, adequately represented?
9. Is the research ethical according to current criteria or, for recent studies, and is there evidence of ethical approval by an appropriate body?
10. Do the conclusions drawn in the research report flow from the analysis, or interpretation, of the data?

If necessary, the authors of the papers will be contacted to request for missing or additional data for clarification. Any disagreements between the reviewers will be resolved through discussion or, if needed, by involving a third reviewer.

The results of the critical appraisal were reported in both narrative and tabular formats.

### Incorporation of Critical Appraisal Results into the Review

All studies were included in the review, regardless of methodological quality. However, studies with low methodological quality should be considered cautiously when interpreting and drawing conclusions from these findings. The details and results of the critical appraisal will be summarized in the main text and provided in full in the supplementary materials.

### Data extraction

Data will be extracted from the studies included in the review by two independent reviewers using the JBI standardized data extraction tool. While the standard tool will be used, any modifications will be described in the text, and the revised version will be appended if necessary.

The extracted data will include the following:

1. Methods for data collection and analysis
2. Country
3. Phenomena of interest
4. Setting/context/culture
5. Participant characteristics and sample size
6. Description of main results

The findings from these studies will be extracted verbatim and assigned a level of credibility as follows:

- Unequivocal: The relationship between the illustration (participant narratives) and findings (research results) is clear and well-supported.

Credible: The relationship between the illustration and findings is not entirely clear, but the results are not completely unreliable.

- Not Supported: No relevant illustration is provided, or the findings are not adequately substantiated.

The following strategies were employed to minimize errors during the data extraction process: Independent data extraction: Two reviewers will independently extract the data.

- Resolution of discrepancies: Any disagreements between the reviewers will be resolved through discussion, and if unresolved, a third reviewer will intervene.

Verification of missing information: If any data are missing or unclear, the authors of the study will be contacted for clarification or additional information.

Transparency in assumptions: Any assumptions made regarding missing or unclear data are explicitly stated in the review.

These measures will ensure the transparency and consistency of the review process.

### Data synthesis

This review employs a meta-aggregation approach to synthesize qualitative research findings using JBI SUMARI. This approach involves the following steps.

#### 1. Collection and Categorization of Findings

The findings extracted from the included studies were gathered and categorized based on their similarities in meaning.

#### 2. Aggregation of Categories

The categorized findings were then further synthesized to generate a comprehensive set of results.

The synthesized findings are presented as a structured set of statements, consolidating the research results into actionable insights for evidence-based practice. These statements will provide guidance on practical applications but will avoid directive language such as “should” or “must.”

### Presentation in Narrative Form

If textual pooling is not feasible, the findings will be presented in narrative format, maintaining the integrity of each study’s results while providing contextual interpretations.

Only findings that meet the following criteria will be included in the synthesis:

- **Unequivocal:** Findings clearly supported by data and well substantiated.
- **Credible:** Findings that are reliable but may lack complete clarity.

Findings that are unclear or lack credibility are excluded from the synthesis to ensure the robustness and reliability of the conclusions.

### Assessing confidence in the findings

In this review, synthesized findings are assessed based on the ConQual approach to establish the level of confidence in qualitative research synthesis. The evaluation results are summarized in the Summary of Findings table, which includes the following elements:

- Type of research
- Dependability score
- Credibility score
- Overall ConQual score

Additionally, the process and criteria for calculating the ConQual score are detailed to ensure transparency in the evaluation. This approach demonstrates the applicability of synthesized qualitative findings to practice and reinforces the reliability of evidence-based decision making.

## Data Availability

None

## Acknowledgements

This review will include acknowledgments of individuals who have made significant contributions. Additionally, it specifies if this review contributes towards a degree award and for which author.

## Declarations

This review emphasizes equity, diversity, and inclusion, with the aim of integrating these values into research. By incorporating the perspectives of authors with experience in nursing practice, the review enhances transparency and reliability, while ensuring a comprehensive synthesis that reflects diverse viewpoints.

## Author contributions

Each author’s specific contributions to this review will be documented as follows: Designing the analysis: Developing an overall analytical framework for the study.

- Providing or collecting data: Gathering relevant literature and data required for the review.

Performing the analysis: Conducting the data analysis and synthesizing the results.

Writing of the manuscript: Drafting and editing sections of the review.

This documentation aimed to clarify each author’s role and ensure transparency in the research process.

## Appendices

Appendix I: Search strategy

In accordance with this protocol, a comprehensive search strategy for at least one electronic database was used to ensure the reproducibility of the review. The full search strategy used in this review is outlined below.

### Database and Search

**Strategy Database:** PubMed (MEDLINE)

### Search Query

((“hematopoietic stem cell transplant nurse” [TIAB] OR “HSCT nurse” [TIAB] OR “bone marrow transplant nurse” [TIAB] OR “stem cell transplant nurse” [TIAB] OR “hematology-oncology nurse” [TIAB] OR “oncology nurse” [TIAB] OR “hematology nurse” [TIAB])

AND

(“Compassion Fatigue” [MeSH] OR compassion fatigue [TIAB] OR “moral distress” [TIAB] OR “psychological stress” [MeSH] OR “coping strategies” [TIAB] OR “coping” [TIAB] OR “professional quality of life” [TIAB] OR “stress levels” [TIAB] OR “difficulty” [TIAB] OR “challenges” [TIAB] OR “burnout” [TIAB] OR “empathy” [TIAB] OR “resilience program*” [TIAB] OR “support program*” [TIAB])

AND

(“hematopoietic stem cell transplant” [TIAB] OR “bone marrow transplant” [TIAB] OR “stem cell transplant” [TIAB] OR “hematologic malignancies” [TIAB] OR “hematology-oncology” [TIAB] OR “oncology” [TIAB] OR “leukemia” [TIAB] OR “blood cancer” [TIAB] OR “hematology” [TIAB] OR “blood disorders” [TIAB])

AND

(“qualitative research” [MeSH] OR “qualitative study” [TIAB] OR “qualitative analysis” [TIAB] OR “mixed-methods” [TIAB] OR “phenomenological study” [TIAB] OR “experiences” [TIAB] OR “thematic analysis” [TIAB]))

**Search Conducted on:** November 25, 2024

This search strategy incorporates a combination of keywords, MeSH terms, and Boolean operators to ensure comprehensive retrieval of relevant studies. This approach was designed to maximize sensitivity and specificity in identifying qualitative research related to the experiences of hematopoietic cell transplant nurses.

## Appendix II: Data extraction instrument

This review will use the standard JBI data extraction tool.

